# Normal values for native T1 at 1.5T in the pericardial fluid of healthy volunteers

**DOI:** 10.1101/2022.08.29.22279326

**Authors:** Simon Thalén, Joao G Ramos, Henrik Engblom, Andreas Sigfridsson, Peder Sörensson, Martin Ugander

## Abstract

**Background:** T1 mapping cardiovascular magnetic resonance (CMR) imaging has been used to characterize pericardial effusions. The aim of this study was to measure pericardial fluid native T1 values in healthy volunteers to establish normal values.

**Methods:** Prospectively recruited volunteers (n=30) underwent CMR at 1.5T and native T1 maps were acquired using a MOLLI 5s(3s)3s acquisition scheme. A volume of pericardial fluid was imaged in a short-axis slice and in a slice perpendicular to the short-axis orientation. A reliable measurement had a region of interest size >10 mm^2^, coefficient of variation <10 %, and a relative difference <5 % between the two slice orientations.

**Results:** In 26/30 (87%) of volunteers, there was a sufficient amount of pericardial fluid to enable reliable measurement. Native T1 of pericardial fluid was 3262±163 (95% normal limits 2943–3581ms), and did not differ in the perpendicular slice orientation (3267±173 ms, p=0.75), due to sex (female 3311±177 vs. male 3220±142 ms, p=0.17), or age (R^2^=0.07, p=0.2)

**Conclusions:** This study shows that T1 values can be reliably measured in the pericardial fluid of healthy volunteers. It is the first to report normal reference ranges for T1 values at 1.5T in the pericardial fluid of healthy volunteers.

## Introduction

Pericardial fluid is a serous ultrafiltrate of plasma normally present between the layers of the pericardium enveloping the heart. An increase in the amount of fluid above approximately 30-35 ml is considered a pericardial effusion [1]. Pericardial effusions can have a wide variety of etiologies, and are a common clinical occurrence. The diagnostic approach includes categorizing the effusion as transudative, due to an imbalance in osmotic and hydrostatic pressures, or exudative, due to an increase in permeability of the pericardium secondary to local inflammation. Biochemical analysis may be performed and interpreted using the Light criteria, originally developed and validated for assessing pleural effusions [2].

With the rapid development of new techniques in cardiovascular magnetic resonance imaging (CMR) there have been several attempts to non-invasively characterize effusion fluids. Early attempts using magnetic field strengths below one tesla were largely unremarkable [3–7]. Later results using diffusion-weighted imaging have shown some promise, but also conflicting results [8–11].

T1-mapping, a technique whereby the T1 value is quantified by imaging on a pixel-by-pixel basis, has now largely superseded the previous T1-weighted methods where the T1 value was encoded into arbitrary signal intensities and only indirectly estimated. T1-mapping CMR has enabled the characterization of focal and global cardiovascular pathologies, and made it possible to reliably and quantitatively perform longitudinal and group comparisons [12].

Non-contrast (native) T1 mapping has successfully been used to classify pericardial and pleural effusions as transudate or exudate [13], and has further been used to successfully differentiate between malignant and non-malignant pleural and ascitic effusions *ex vivo* [14]. The rationale for these approaches is that proteins and cells leaking through a permeable inflamed pericardium and into the pericardial fluid have paramagnetic properties that lower the T1 value of the effusion fluid. Consequently, a low native T1 value would be consistent with an exudate and a high value with a transudate.

The current study aimed to measure T1 in pericardial fluid in healthy volunteers by determining the feasibility of these measurements and establishing normal values.

## Methods

### Volunteers

Healthy volunteers (n=30) were recruited. Exclusion criteria were any known cardiovascular pathology or metallic implants not safe for MRI.

### Image acquisition

T1 maps were acquired at end diastole using a modified look-locker inversion recovery (MOLLI) sequence [16] at 1.5T (MAGNETOM Aera, Siemens Healthcare, Erlangen, Germany) using motion-correction [17]. A 5s(3s)3s acquisition scheme where two inversions were made and images acquired once per heartbeat during five seconds after the first inversion, followed by a three-second pause, and images acquired once per heartbeat during three seconds after the second inversion [18]. Typical image acquisition parameters were: flip angle 35°, matrix size 256 × 136–158, slice thickness 8 mm, initial inversion time 129 ms, field of view 300-410 × 241–384 mm^2^, parallel imaging factor 2.

A short-axis (SA) stack of T1-maps encompassing the heart and the ascending aorta was acquired and screened for the presence of pericardial fluid visible as areas with native T1 values above 2500 ms. A slice perpendicular (P) to the short-axis (SA) slice was then prescribed to enable two perpendicular measurements.

### Image analysis

Image analysis was performed on clinical workstations using clinical imaging software (syngo.via VB30A; Siemens Healthcare GmbH, Erlangen, Germany). Regions of interest (ROI) were manually prescribed in the same volume of pericardial fluid of both the SA and P slices. A three-dimensional cursor feature was used to ensure measurement in the same volume of pericardial fluid in both slice orientations.

To evaluate measurement reliability, the following criteria were used to define a reliable measurement: 1) the ROI size needed to be greater than 10 mm^2^, 2) the coefficient of variation of measured values within the ROI needed to be less than 10%, and 3) the relative difference in measured values between the SA and P slice orientations needed to be below 5%. The coefficient of variation was defined as the ratio of the standard deviation to the mean. The relative difference in native T1 between SA and P measurements was defined as |SA-P| / [(SA+P)/2].

### Statistical analysis

The normality of distributions was assessed both visually using Q-Q plots and the Shapiro-Wilks test and data reported as median [interquartile range] or mean±SD as appropriate. Mean value comparisons were made using parametric paired or unpaired t-tests as appropriate. A p-value of less than 0.05 was considered statistically significant. Normal reference ranges were calculated as mean ± 1.96*standard deviation. Statistical calculations were performed using the software package R (R Core Team 2020, Vienna, Austria).

## Results

The healthy volunteers averaged 35 (range 20–61) years of age, 14 (46 %) were female and 26/30 (87 %) had a sufficient amount of pericardial fluid to enable reliable measurement. The characteristics of the volunteers are shown in table 1 and an example T1 map from one volunteer is shown in figure 1.

**Table 1.** Baseline variables for healthy volunteers.

| Characteristic |  |
| --- | --- |
| Reliably measurable T1, n/n (%) | 26/30 (87) |
| Female sex, n/n (%) | 12/26 (46) |
| Age, years | 35 [46-65] |
| Height, cm | 174±10 |
| Weight, kg | 71±12 |
| Body mass index, kg/m<sup>2</sup> | 22±2 |
| Body surface area, m<sup>2</sup> | 1.8±0.2 |

Data are presented as numbers (n) and percent, mean±SD, or median [interquartile range].

**Figure 1.** An example of native T1 maps taken in short axis (left) and perpendicular (right) slice orientations.

Native T1 of pericardial fluid was 3262±163 (95% normal limits 2943–3581ms), and did not differ in the perpendicular slice orientation (3267±173 ms, p=0.75). Native T1 of pericardial fluid did not differ due to sex (female 3311±177 vs. male 3220±142 ms, p=0.17), age (R^2^=0.07, p=0.2) or heart rate (R^2^=0.07, p=0.18).

## Discussion

This study shows that T1 values can be reliably measured in the pericardial fluid of healthy volunteers. It is the first to report normal reference ranges for T1 values in the pericardial fluid of healthy volunteers. Normal values are of importance as the pericardial fluid of healthy volunteers is usually not available for biochemical analysis.

The native T1 values of the pericardial fluid in the healthy volunteers in the present study are comparable to native T1 values in pericardial effusions presumed to be transudate [13]. When applying the native T1 cut-off value proposed for exudates in that study (3105 ms), two healthy volunteers from the present study had a lower value.

In one study, 0.6 % of CMR exams were found to have incidental findings of clinically significant pericardial effusion [19]. A non-invasive alternative to diagnostic pericardiocentesis would have clinical significance as all invasive procedures involve some degree of risk. International guidelines recommend pericardiocentesis with biochemical testing upon suspicion of bacterial or neoplastic pericardial effusion [20]. A non-invasive method for characterizing pericardial fluid as transudate, and hence not of bacterial or neoplastic etiology, could then be used to avoid unnecessary diagnostic pericardiocentesis. If on the other hand a pericardial effusion is characterized as an exudate, CMR imaging provides detailed anatomical information highly relevant to the pericardiocentesis procedure itself.

## Limitations

A potential limitation of the current study is the size of ROI required to measure the small volumes of normal pericardial fluid present in healthy volunteers. The matrix size, field of view and slice thickness used in this study are equal to those in routine clinical use and translate to voxel dimensions of approximately 1.5 × 1.5 × 6 mm or 13.5 mm^3^. The median ROI size of 18 mm^2^ translates to 8 in-plane voxels, or image pixels, per ROI whereas international CMR consensus guidelines recommend avoiding ROI sizes below 20 image pixels for T1 mapping of the myocardium [21]. Even though some partial volume is inevitable, we attempted to overcome this limitation by employing a minimum ROI size criteria, a within-ROI variance criteria, and a between-ROI variance criteria with separately acquired and perpendicularly oriented measurements. There are currently no studies or guidelines available to inform the choice, or specific values, of the above criteria for measurements of T1 in pericardial fluid. They were chosen for purposes of consistency based on clinical judgement.

The sequence used in this study was the 5s(3s)3s variant of MOLLI. Although, other cardiac T1 mapping sequences are available such as SASHA [22], shMOLLI [23], SAPPHIRE [24] among others, MOLLI was chosen as it is the cardiac T1 mapping sequence most widely used clinically. The 5(3)3 variant of MOLLI sequence has been shown to be heart-rate dependent, while the 5s(3s)3s is an improvement, some remaining heart rate dependence is to be expected [25]. The error introduced at high heart rates is due to incomplete relaxation and residual longitudinal magnetization at the time of the next inversion, leading to a lower measured T1 value. The amount of residual longitudinal magnetization is not only increased at high heart rates but also at higher T1 values. Notably, the native T1 values for pericardial fluid in the current study did not vary with heart rate. The T1 values of the pericardial fluid of healthy volunteers measured in this study are considerably higher than the proposed validated range for measuring native T1 (below 1200 ms) [25]. An underestimation in T1 values is thus to be expected, and the normal values presented in this study should not be generalized beyond the specific 5s(3s)3s variant of the MOLLI sequence. To our knowledge there are no simulation, phantom or volunteer studies that investigate the accuracy and precision of cardiac T1 mapping sequences for the higher T1 values of serous fluids.

## Conclusions

T1 can be reliably measured at 1.5T in a normal volume of pericardial fluid in healthy volunteers, and normal reference values are presented. The use of native T1 mapping characterize pericardial fluid is currently in its infancy and further research is required to determine the value of this approach. The precision and accuracy of specific cardiac T1 mapping sequences for measurement of the higher T1 values of serous fluids is an open question.

## Data Availability

All data produced in the present study are available upon reasonable request to the authors

## Declarations

### Ethics approval and consent to participate

The study was approved by the Swedish Ethical Review Authority (application number 2011/1077-31/3) and conducted according to the principles of the Declaration of Helsinki. All participants were enrolled following written informed consent.

### Consent for publication

Written informed consent was obtained from patients for publication of their individual details on a group level and anonymized images in this manuscript. The consent form is held in the patients’ clinical notes and is available for review by the Editor-in-Chief.

### Availability of data and materials

The datasets analysed in the current study are available from the corresponding author upon reasonable request.

### Author contributions

ST contributed to design of the study, performed data acquisition, data analysis and wrote the manuscript. AS supervised the technical aspects of data acquisition. JGR, HE and PS participated in data acquisition and all authors substantially contributed to the interpretation of the results. MU designed the study, contributed to writing of the manuscript, and is overall guarantor of the scientific integrity of the study.

## Acknowledgements

We acknowledge the support provided by the CMR technologists at the Department of Clinical Physiology, Karolinska University Hospital. Karolinska University Hospital has a research and development agreement regarding CMR with Siemens Healthineers.

